# A SEIR-like model with a time-dependent contagion factor describes the dynamics of the Covid-19 pandemic

**DOI:** 10.1101/2020.08.06.20169557

**Authors:** Ronald Dickman

## Abstract

I consider a simple, deterministic SEIR-like model without spatial or age structure, including a presymptomatic state and distinguishing between reported and nonreported infected individuals. Using a time-dependent contagion factor *β*(*t*) (in the form a piecewise constant function) and literature values for other epidemiological parameters, I obtain good fits to observational data for the cumulative number of confirmed cases in over 160 regions (103 countries, 24 Brazilian states and 34 U.S. counties). The evolution of *β* is useful for characterizing the state of the epidemic. The analysis provides insight into general trends associated with the pandemic, such as the tendency toward reduced contagion, and the fraction of the population exposed to the virus.

## I. INTRODUCTION

The Covid-19 pandemic has stimulated the development of epidemic modelling in various directions: identification of epidemiological parameters, projection of general trends under different intervention policies, analysis of the propagation of outbreaks between geographic regions, age and/or social classes, and so on. In this context it is desirable to deploy a range of models, with varied levels of detail. Highly detailed models afford greater realism, but at the cost of a proliferation of unknown parameters. In the present study I analyze a simple, deterministic model without spatial or age-class structure. The question motivating this study is whether a plausible model with a limited number of free parameters can reproduce the broad features of the evolution of outbreaks. A related effort, using a discrete-time stochastic model, has been developed by Karlen [1], while Paiva et al. test the predictive power of such an approach [2]. Further applications of simple deterministic models to the current pandemic may be found in [3, 4].

The simplest deterministic, dynamic epidemic models belong to the SIR family (SEIR, SEAIR, etc.) [5–7]. The choice of the specific variant from this class is guided by certain characteristics of the Covid-19 epidemic: (i) a delay between exposure to the virus and becoming cantagious; (ii) a further delay between becoming contagious and exhibiting symptoms; (iii) the broad range of symptoms, ranging from negligible to life-threatening; and (iv) significant undercounting of cases, due in part to the large variation in gravity of cases as well as to limited testing capacity. These observations suggest that the minimal sequence of states (susceptible, infected, removed) be expanded to include exposed and presymptomatic states between S and I, and further, that the class of infecteds be split into confirmed and unconfirmed subclasses.

An essential fact regarding the present state of the pandemic is that the fraction of exposed individuals is globally very small, with most estimates suggesting just a few percent, far smaller than what would be necessary to slow the growth of outbreaks through the depletion of susceptibles. (In exceptional, resticted regions, such as New York City, for which some studies suggest that ~ 20% of the population may have been exposed [8], there may be significant reduction in the suscpetible fraction, albeit still below the level required for herd immunity.) Thus the reduction of case-number growth rates in almost all outbreaks (at least initially; see Fig. 1) cannot be attributed to large-scale exposure and immunity.

**FIG. 1:**
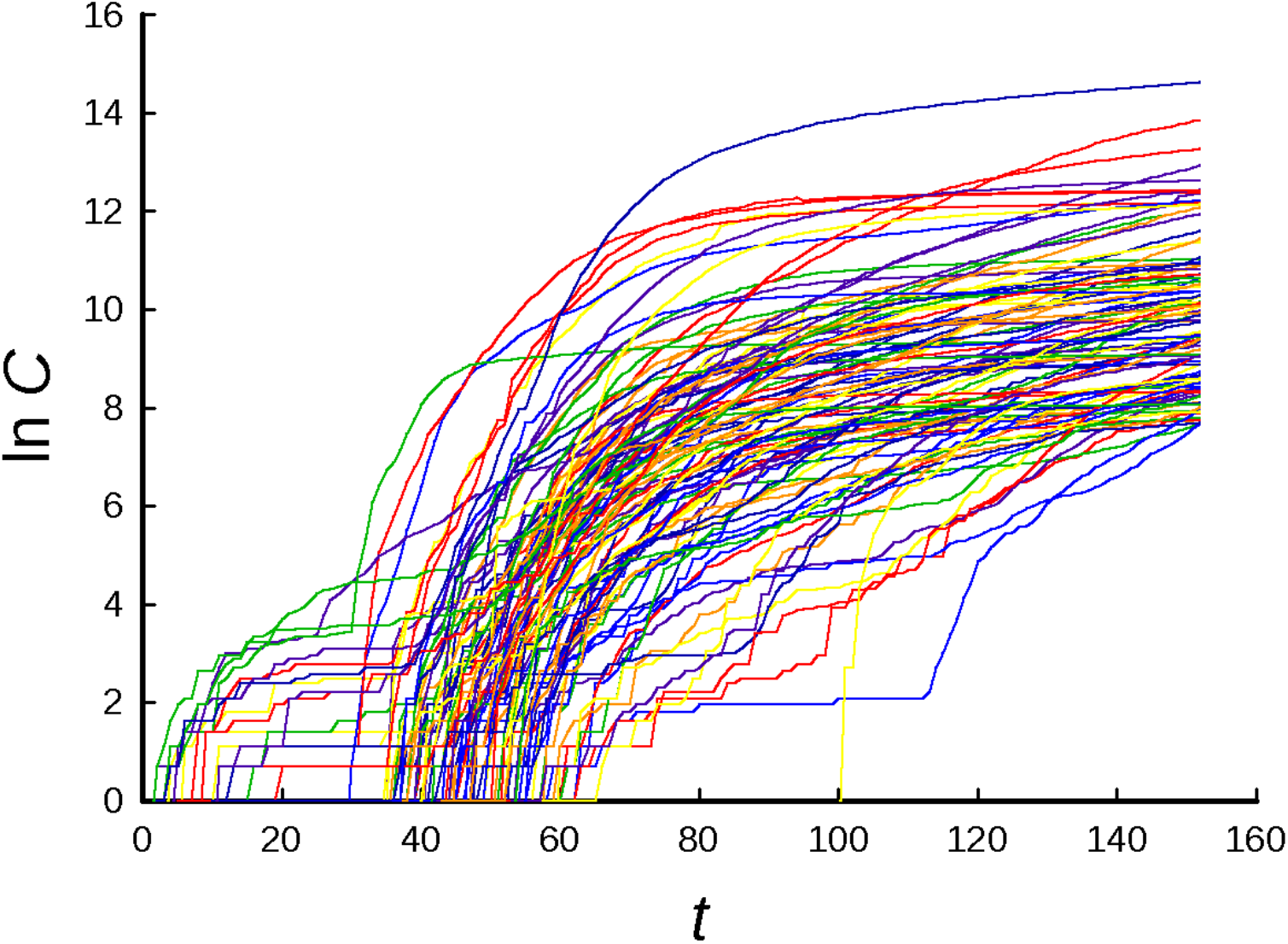
Evolution of cumulative case numbers in 103 regions (countries and large provinces) with at least 2000 reported cases as of 20 June 2020. Data from [11]

The dominant factor in cutting growth rates is mitigation or intervention in the form of social distancing, mask use, reduced mobility, lockdown, testing and tracking, etc. The effect of such measures, broadly speaking, is to reduce contagion. It is therefore essential to include a time-dependent contagion parameter in the model. (The fact that the adoption of, and adherence to, mitigations are unpredictible political and social events highlights the difficulty of long-term forecasting using epidemic models. One may nevertheless hope that such models can suggest the likely consequences of different mitigation policies.)

In the model adopted here, a susceptible individual may become exposed through contact with a presymptomatic or infected individual at a rate propotional to the contagion parameter, *β*(*t*), which in fact contains all of the adjustable parameters of the model. (Other epidemiological parameters are fixed at values taken from the literature.) In keeping with the minimalist approach adopted here, the functional representation of *β* is close to the simplest possible. It is worth noting that *β* is linearly proportional to the basic reproduction number *R*_0_, and can be related to the relative growth rate *α*. The history of *β*(*t*), in a given region or globally, obtained by fitting the data for cumulative case numbers, affords some measure of the control (or resurgence) of the epidemic, and allows to detect some overall trends, which appear to hold idependently of region size or location. I find that a rather simple (piecewise constant) representation yields reasonable fits to most of the time series.

The remainder of this paper is organized as follows. In Sec. II I define the model and discuss its behavior in the linear regime, as well as presenting some examples of the response to changes in *β*. In Sec. III, model fits to 103 countries, 24 Brazilian states, and 34 U.S. counties and cities with large outbreaks are presented. Simulations of a *stochastic* version of the model are used to characterize uncertainties in *β*(*t*), and the sensitivity of the quality of fit to variations in *β*(*t*) is also examined. Sec. IV contains a discussion of overall trends on contagion rates and fractions of susceptibles, as well as examining the sensitivity of *β*(*t*) to changes in the epidemiological parameters. I close in Sec. V with a summary and comments on possible extensions of this study.

## II. SEAUCR MODEL

The model divides the population into the following categories: susceptible (S); exposed (E); presymptomatic (A); infected but unconfirmed (U); infected and confirmed (C); removed (R). It follows the general lines of the model used by Costa, Cota and Ferreira [9] to represent a given region within a network. (In the present study, however, there is no network or other spatial structure.) The states and allowed transitions are shown in Fig. 2. Denoting the fraction of the population in a given state by the corresponding lower-case letter, we have the following set of equations:

**FIG. 2:**
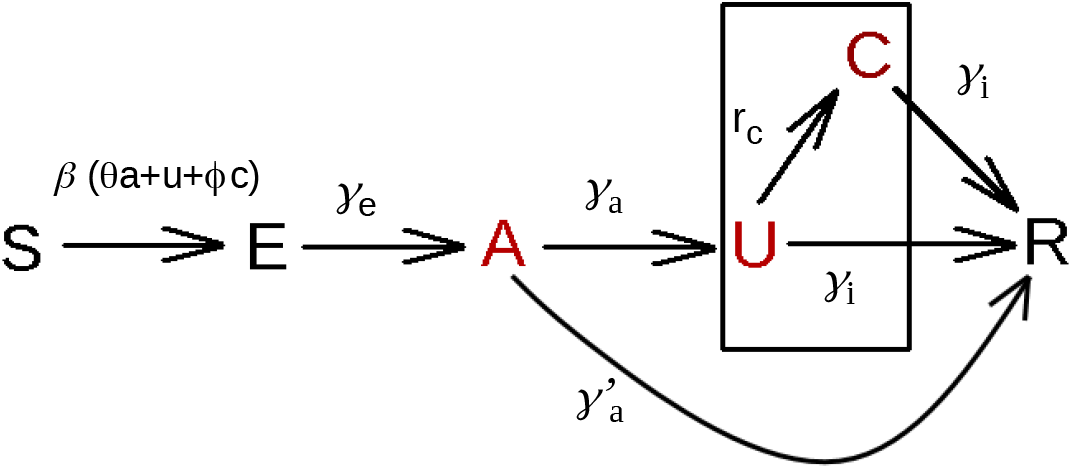
States and transitions. The only distinction between substates U and C is that the former contains cases that have not been reported, whereas those in the latter class have been.

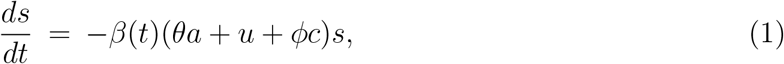

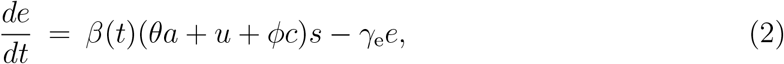

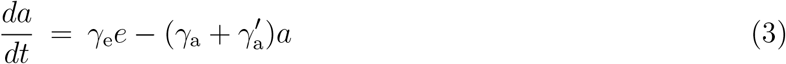

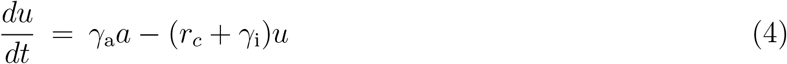

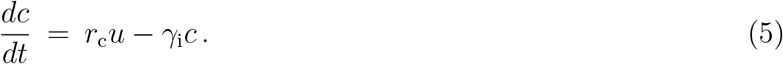

Of central importance in this study is the cumulative fraction of confirmed cases, *C*(*t*), which is governed by,

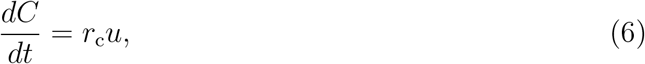

with the initial value, *C*(0), estimated from observational data, as discussed below.

I adopt the following set of fixed-parameter values, used in [9, 10]: *γ*_e_ = *γ*_a_ = (2.6 day)^−1^, 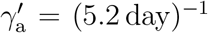, γ_i_ = (3.2 day)^−1^, *r_c_* = (10 day)^−1^, *ϕ* = 0.3 and *θ* =1. The choice *ϕ* < 1 reflects the hypothesis that a fraction of individuals with confirmed infections are isolated (or self-isolate) from the general population. (Although I use *θ* = 1 in the present analysis, a value smaller than unity could be used to represent a reduced level of contagion during the presymptomatic phase.) I regard these parameter values as plausible. Precise values are not available, and in any case a realistic treatment of disease progression would use nontrivial distributions of transition times (leading to integrodifferential equations), rather than the exponential distributions implicit in SIR-like models. It seems likely that in fitting data, modest changes in the parameter values can be compensated by rescaling *β* (see Sec. IV.B).

The fixed parameters determine the response times of the epidemic as well as the fraction of cases that are reported. From Fig. 2, one sees that to be reported, a presymptomatic individual must progress to state U (and not directly to state R), and thence to C (rather than directly to R), so that the fraction of reported cases is:

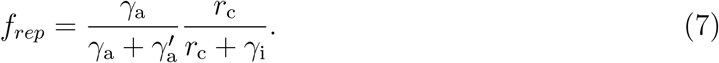

The first factor is connected to exposure leading to identifiable symptoms, while the second involves severity of symptoms as well as testing capacity. Both are subject to change as testing policies and capacities are altered. For the parameter values used here, *f*_rep_ ≃ 0.207. In the studies discussed below, the model equations are integrated numerically using a fourth-order Runge-Kutta scheme with a timestep of 0.1 [12]. All studies use *θ* =1. i.e., unreported infected and presymptomatic individuals are considered equally contagious.

### A. Initial-stage evolution

Equations (1-5) become a linear set when we fix *s* = *s*_0_, as holds (with *s*_0_ = 1) in the initial phase of an outbreak. Setting *β*(*t*) = *β* (independent of time), the equations for *e, a*, *u* and *c* admit an exponential solution, with *e* = *ɛe^αt^*, and similarly for the other variables. Substituting the exponential forms into the linear equations (and setting *s*_0_ = 1), one finds,

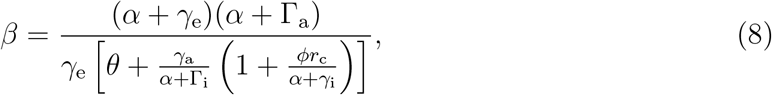

where 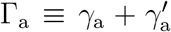 and Γ_i_ = γ_i_ + *r_c_*. The amplitudes in the exponential-growth regime follow,

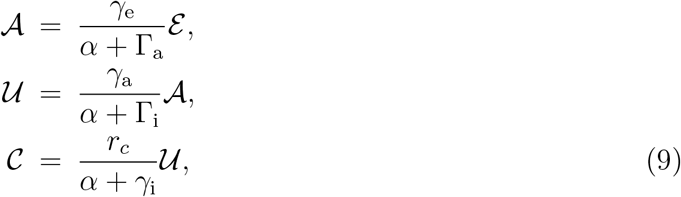

while the cumulative number of reported cases is *C*(*t*) = *Ƶ*(*e^αt^* − 1), well approximated by *Ƶe^αt^* for *αt* ≫ 1, with 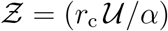.

Note that in practice, we only have access to the daily number of confirmed cases and its accumulated value, *C*(*t*). To determine the initial conditions for integrating the model equations, we localize the earliest apparent linear-growth period in the graph of ln *C* versus *t*, (always discarding points with ln *C* < 2), and fit a straight line to these data, yielding *α* and *Ƶ*. We use these values to fix the other amplitudes and the initial value of *β*(*t*), denoted *β*_1_ in what follows.

Analysis of the linear regime also furnishes an expression for the basic reproduction number *R*_0_ in terms of the other parameters. Consider the initial condition *e* = *e*_0_ = 1/*N* and *a* = *u* = *c* =0, i.e., one exposed in a population of *N −* 1 ≃ *N* susceptibles. The mean number of exposed individuals at time *t* is 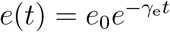, so that the mean number of presymptomatics, due to the single initial exposed individual, is

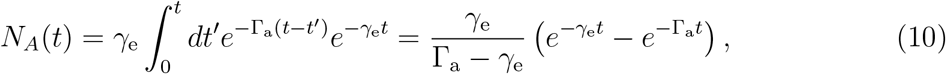

yielding a mean number of new exposed individuals,

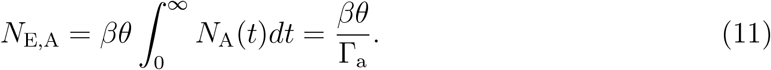

Treating the contribtions due to states U and C in an analogous manner, we arrive at the mean number of new exposed individuals that are down to the initial one:

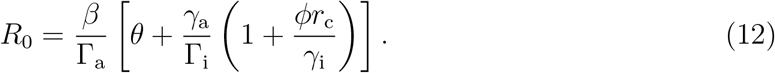

Using Eq. (8), one verifies that *α* = 0 corresponds to *R*_0_ = 1, as expected. For the parameters used here *α* = 0 corresponds to *β* = 0.28534 ≡ *β*_c_. In what follows, the relative growth rate of exposed individuals, 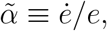, is a convenient index of the state of the epidemic.

### B. Time-dependent contagion parameter

Given an observational time series for the cumulative case number *C*(*t*) of the form shown in Fig. 1, we can estimate the initial exponential growth rate, *α*_1_, and calculate the initial value of *β* using Eq. (8). Reproducing the subsequent evolution requires determining the function *β*(*t*) over the available time interval. I adopt a simple functional form for *β*(*t*) incorporating a small set of adjustable parameters to be determined by least-squares fitting. Consider a contagion factor that switches between *β*_1_ and *β*_2_ during the interval [*τ*, *τ*′], and is constant outside this interval:

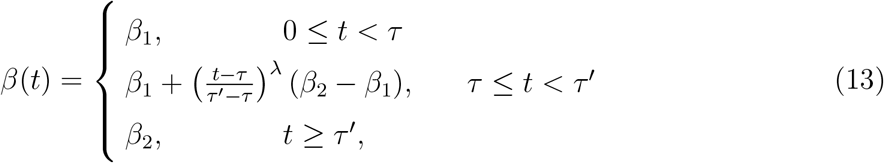

with λ ≥ 0. Fits to data using this expression lead, in general, to best-fit values of λ near zero, suggesting the use of a step function for *β*(*t*). I therefore adopt the following piecewise constant expression,

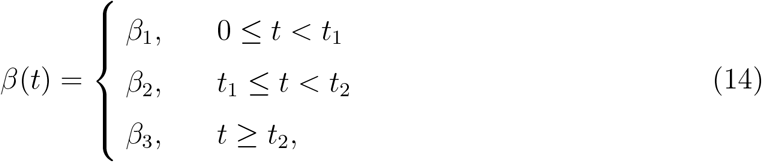

Recalling that *β*_1_ has already been fixed in the analysis of the initial exponential-growth phase, there are four free parameters (two switching times, and two additional *β* values) available to fit the data for *t* ≥ *t*_1_. Denoting the difference between the model and observational values of ln C at day *j* by Δ*_j_*, the objective is to vary the four free parameters simultaneously, so as to minimize the weighted square error,

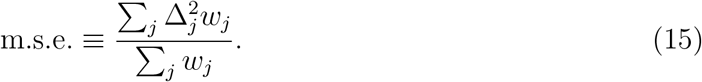

Two weighting functions are used: *w_j_* = *j*^2^ (for overall fits), and *w_j_* = exp[*j*/10] for situations in which one requires more precise parameter estimates for the final portion of the time series.

Before turning to analyses of observational time series, it is interesting to examine the effect of a changing *β* on the model evolution. Figures 3, 4 and 5 show evolutions in which *β* is reduced from a supercritical value to another, smaller supercritical value; from a supercritical to a subcritical value; and from a supercritical to a larger value. The curves for *e*(*t*) show the expected discontinuity in slope, while those for *a, u* and *c* appear smooth, although they have discontinuities in progressively higher derivatives. Of note is the rapid switch between exponential regimes in the curves for *e*, somewhat longer delays for *a, u* and *c*, and much longer delays for *C*(*t*) to fully enter the new exponential growth regime (or to attain a constant value for a subcritical *β* value). This observation no doubt underlies the utility of a *discontinuous β*(*t*) in fitting cumulative case numbers. Another important conclusion is that a subcritical contagion rate (*β* < *β*_c_) does not immediately manifest itself as a flat *C*(*t*); there may be a delay of several weeks.

**FIG. 3:**
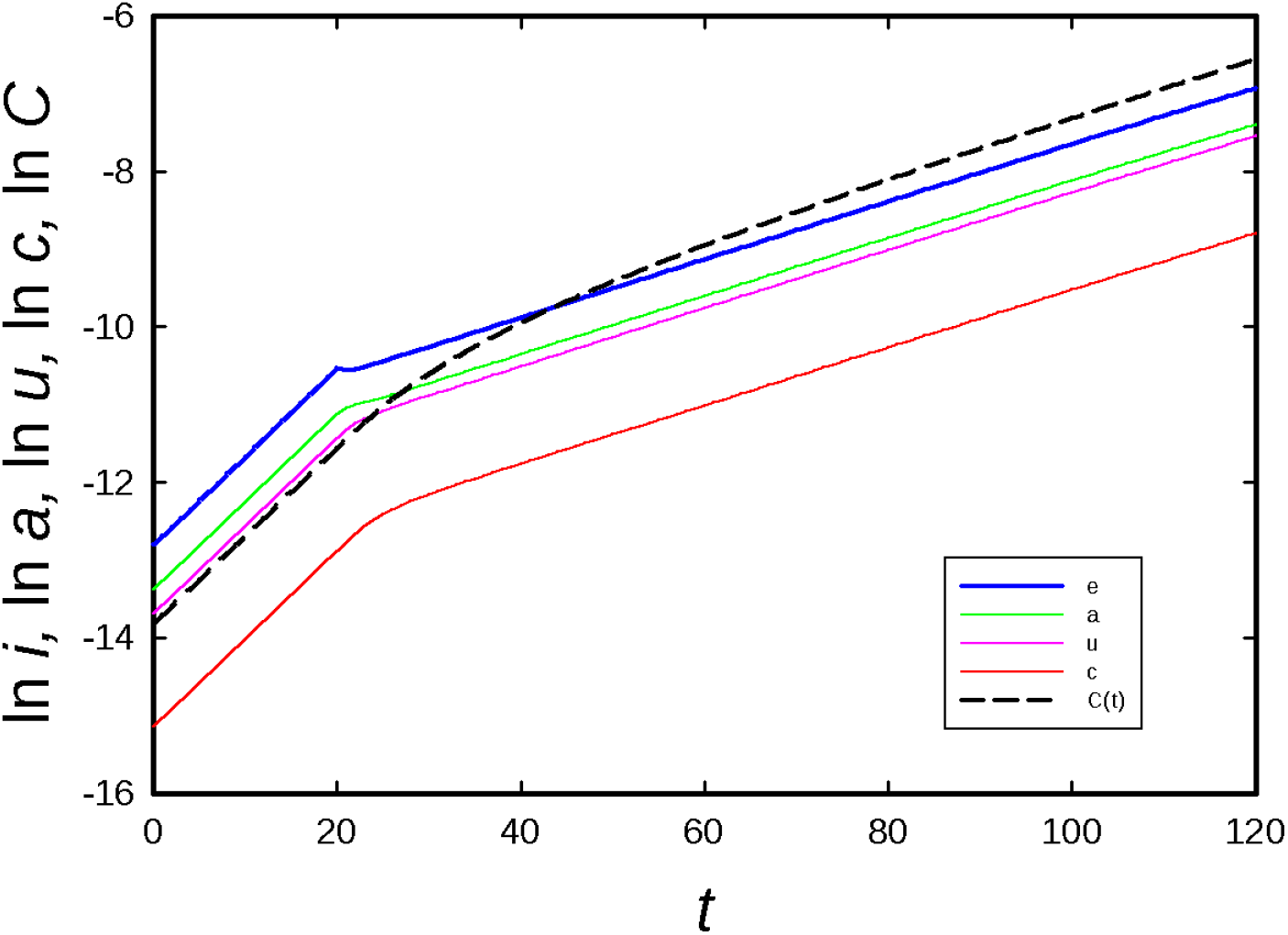
Model evolution starting in the exponential-growth regime with *β* = 0.5 for *t <* 20, with *β* reduced to 0.35 for *t* ≥ 20. The delay times for *e, a, u, c* and *C* to attain their new exponential growth regimes to with 2% are 2, 2, 4, 10, and 65 days, respectively.

**FIG. 4:**
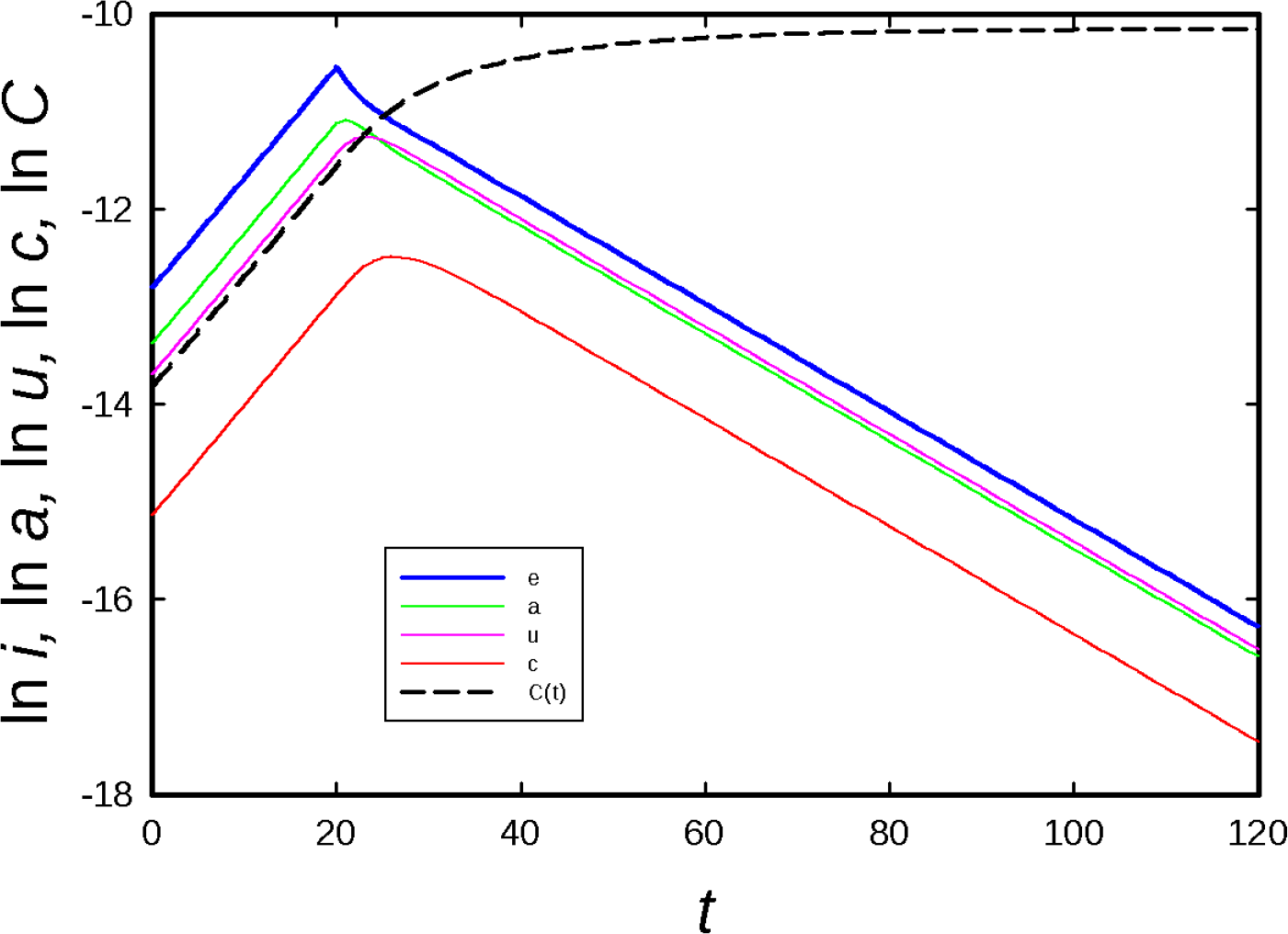
Model evolution starting in the exponential-growth regime with *β* = 0.5 for *t <* 20, with *β* reduced to 0.203 < *β*_c_ for *t* ≥ 20. Delay times for *e, a, u, c* and *C* to attain their new asymptotic behaviors to within 2% are 4, 6, 6, 15, and 63 days, respectively.

**FIG. 5:**
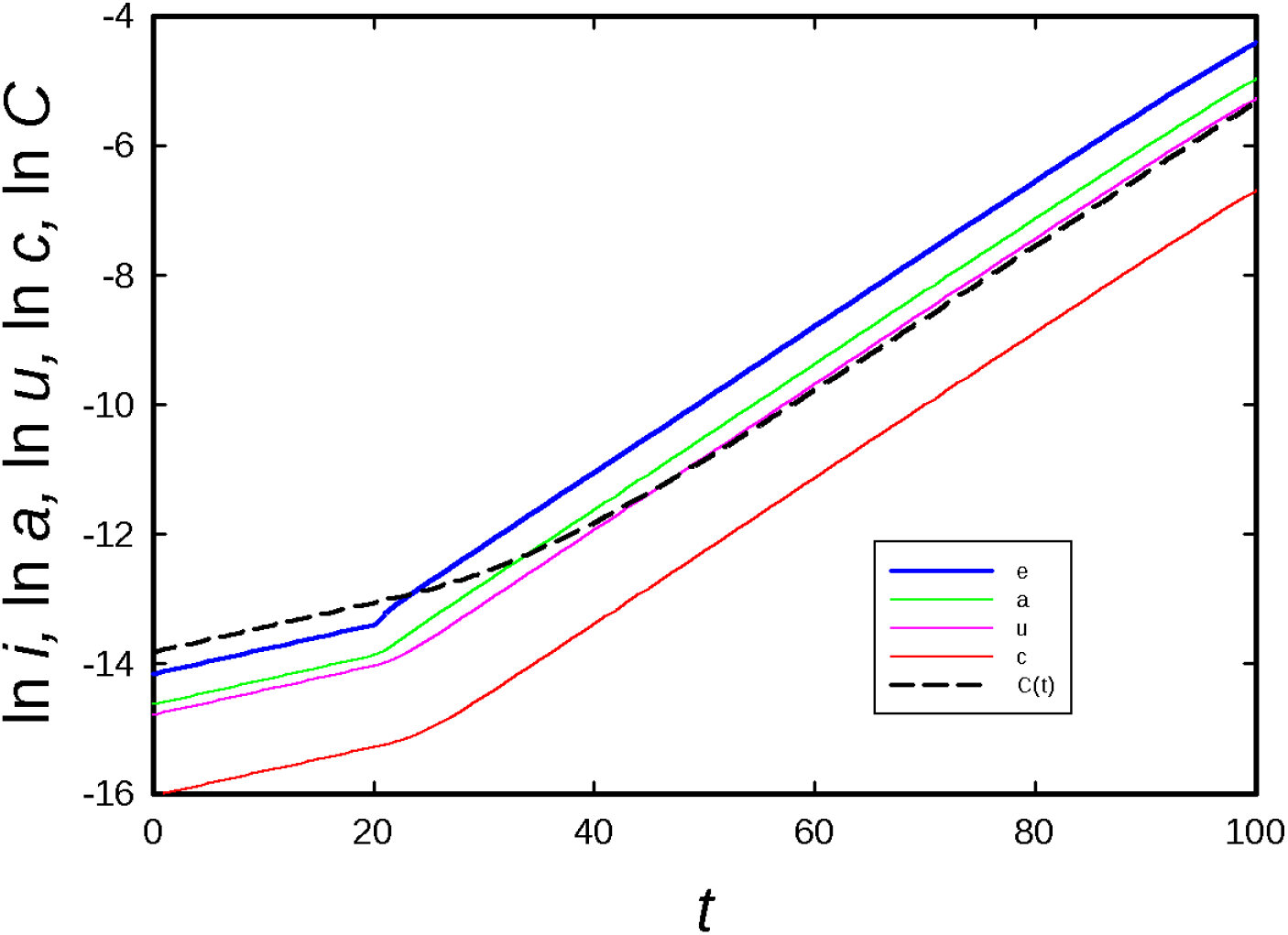
Model evolution starting in the exponential-growth regime with *β* = 0.35 for *t* < 20, with *β* increased to 0.5 for *t* ≥ 20. Delay times for *e, a, u, c* and *C* to attain their new regimes to within 2% are 2, 2, 4, 8, and 38 days, respectively.

## III. RESULTS

In this section I present results of fitting the model parameters to the cumulative numbers of reported cases. Figure 6 shows a typical example, illustrating the choice of the initial exponential-growth regime. The piecewise-constant function for *β*(*t*), taking three distinct values, captures the principal features of the time series, but not the minor fluctuations or (possibly spurious) small jumps near days 22 and 28. Visually, the quality of fit is typical of most cases although, as noted below, there are exceptions. In the following subsections I review the results for U.S. counties and cities, Brazilian states, and countries. Given the somewhat noisy character of the time series, I opt for resolutions of 0.01 in *β*_2_ and *β*_3_, and one day in *t*_1_ and *t*_2_.

**FIG. 6:**
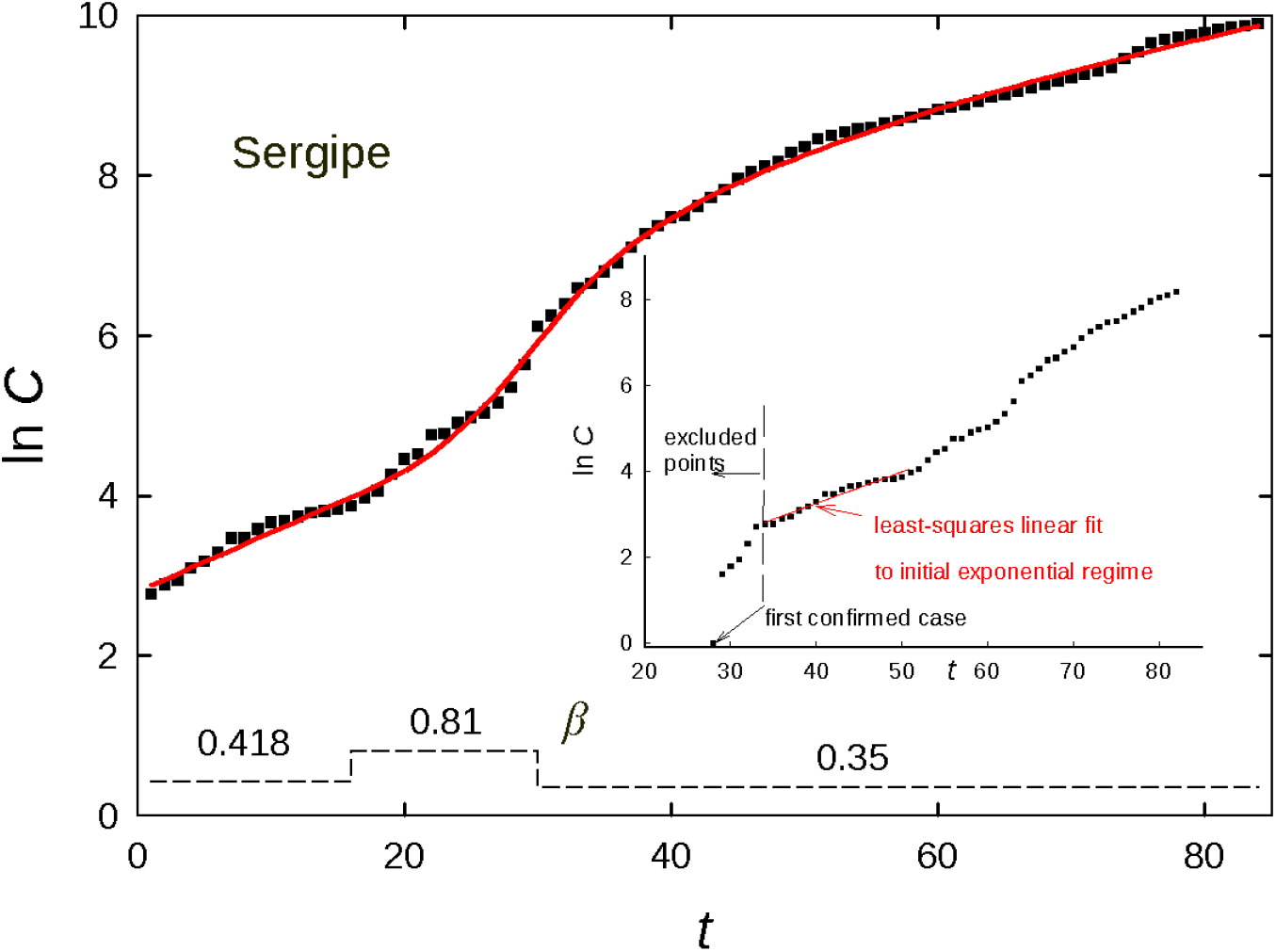
Data (points) and fit (curve) for cumulative case number in the Brazilian state of Sergipe up to 23 June 2020. The dashed line shows *β*(*t*) fit to these data, with numerical values above. Weight function *w_t_* = *t*^2^. Inset: Raw data, showing the initial exponential-growth regime chosen for linear fitting, and excluded points prior to this regime.

### A. U.S. counties

I fit data from the 34 U.S. counties or large cities that had 10 000 or more confirmed cases as of 12 June 2020 [11]. Most of the fits are of good quality, as illustrated by that for Cook Co., Illinois; a few are of poorer quality as exemplified by Nassau Co., New York (see Fig. 7). In the latter case a fourth time interval (and an associated *β* value) would be required to fit the full time series. An interesting point regarding the Cook Co. time series is that although the final *β* value is 0.30, slightly above *β_c_*, the epidemic is in fact becoming smaller (i.e., 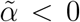 in the final part of the time series). This is because the final susceptible fraction *s_f_* ≃ 0.88, so that the effective contagion rate, *β*_3_*s*, is about 0.26. Note that this conclusion depends on the fraction of reported cases, which is 0.207 for the parameters employed here. The full set of fits to 34 regions is shown in the video: tmser12-6a

**FIG. 7:**
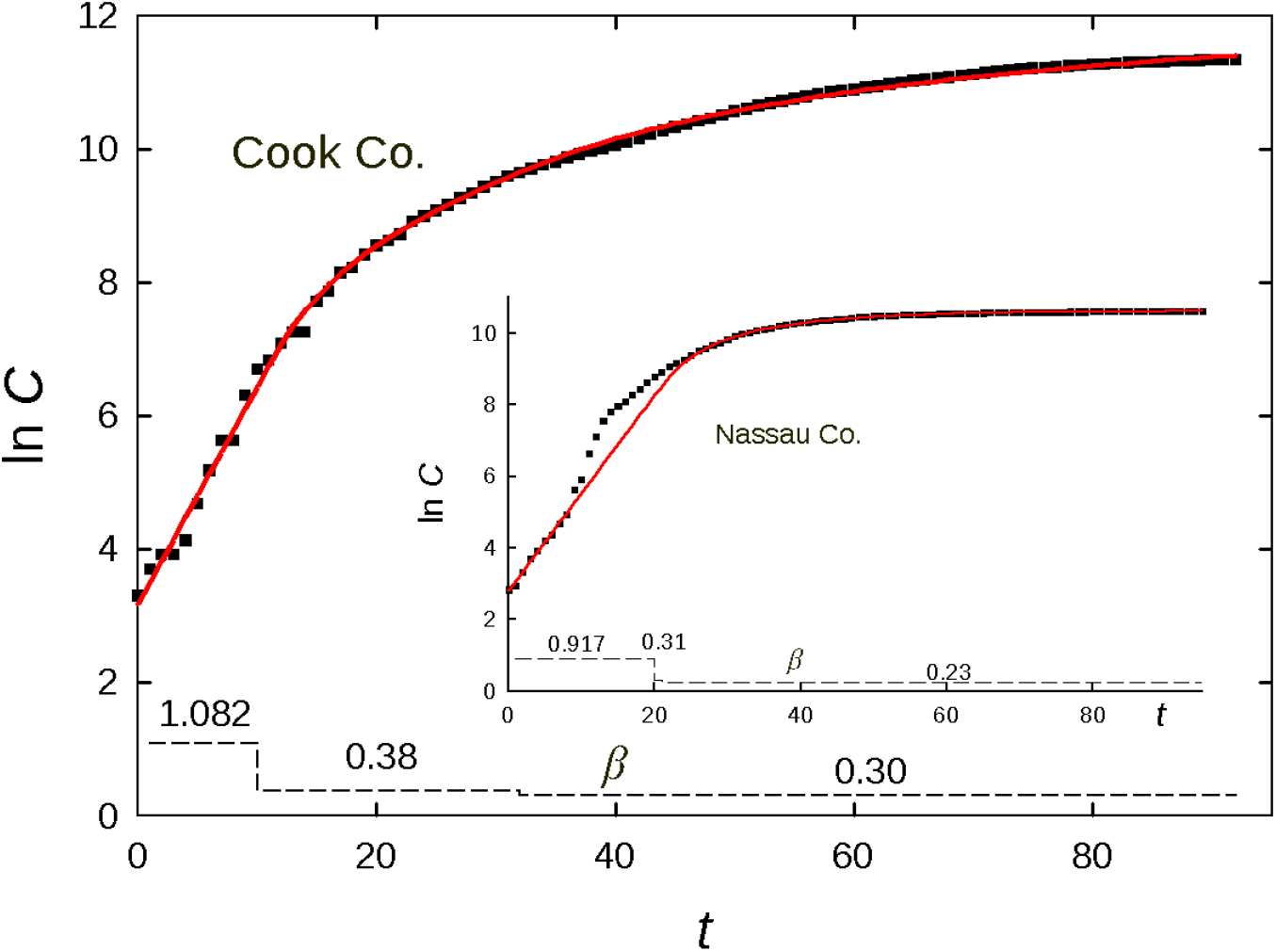
Data (points) and fits (curves) for cumulative case numbers in Cook Co., Illinois (main graph) and Nassau Co., New York (inset) up to 12 June 2020. The dashed lines show *β*(*t*) fit to these data, with numerical values above. Weight function *w_t_* = *t*^2^.

It is instructive to compare the effect of chaging the weight from *t*^2^ to an exponentially growing function, exp[*t*/10]. The comparison for Maricopa Co., Arizona shows that the latter weight yields a better fit to the final two weeks or so, at the cost of of somewhat larger errors for the intermediate period (see Fig. 8). The r.m.s. errors of the two fits are approximately 0.07 (*t*^2^ weighting) and 0.10 (exponential weighting).

**FIG. 8:**
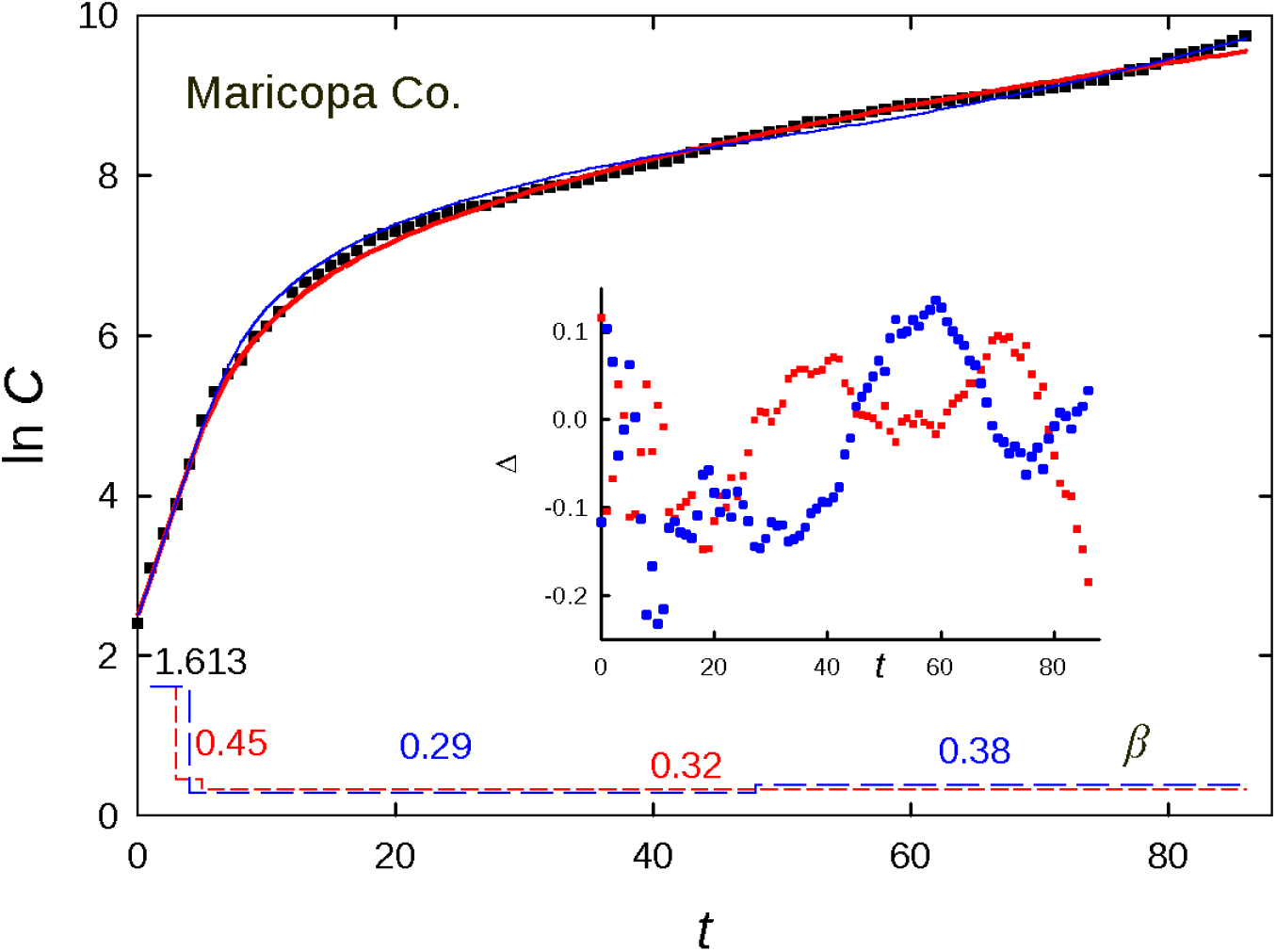
Data (points) and fits (curves) for cumulative case numbers in Maricopa Co., Arizona up to 12 June 2020. The red curve is obtained using the weight function *w_t_* = *t*^2^, the blue using *w_t_* = exp[*t*/10]. The dashed lines show the corresponding β(*t*), with numerical values above, following the same color code. Inset: Residuals Δ for the two fits.

### B. Brazilian states

I fit data from 23 Brazilian states and the Distrito Federal (up to 24 June 2020), that had 2 000 or more confirmed cases as of 2 June 2020 [13]. A typical example is shown in Fig. 6; the full set of fits is shown in the video: tmserst24-6

### C. Countries

Fits to the data [11] for 103 regions (countries or in some cases, provinces) are performed in a similar manner. The set includes those regions having at least 2 000 confirmed cases up to 20 June 2020, excluding China. The full set is shown here: tmser20-6. In about one fifth of the regions studied, the fit appears to miss some portion of the data in a manner that cannot be attributed to an erratic time series, although the overall trends are generally captured; an additional interval seems to be required in these cases. In the “typical” scenario of a smoothly varying curvature, three *β* values appear to be sufficient. Several examples are shown in Fig. 9. The fits for Azerbaijan, Costa Rica, Japan and Thailand are quite good. The data for Ecuador are pathological (discontinuous and nonmonotonic), while those for Senegal suggest the appearance of a second outbreak at some moment prior to day 30.

**FIG. 9:**
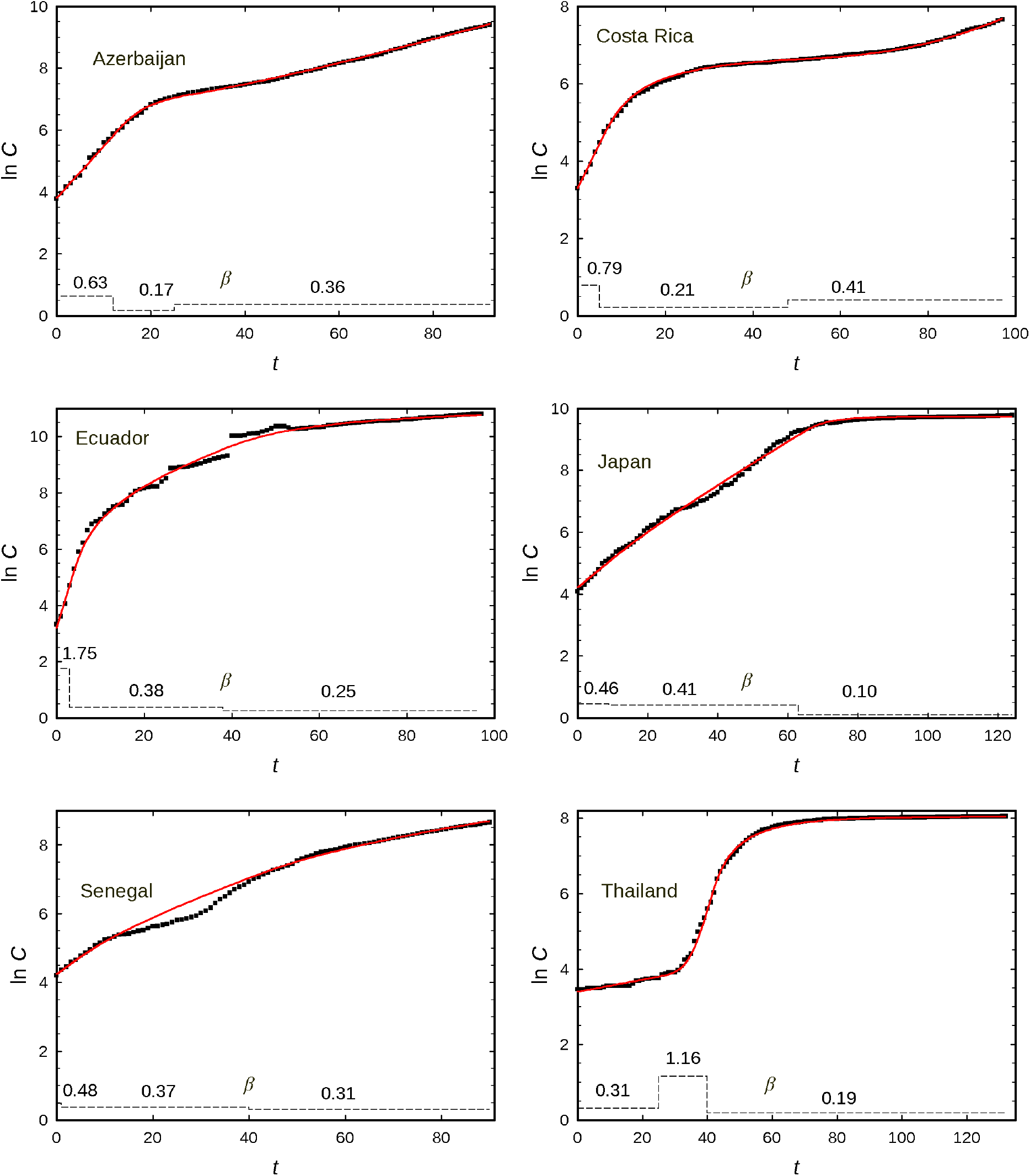
Fits to data for cumulative case numbers in six countries up to 20 June 2020. Weight function *w_t_* = *t*^2^.

### D. Stochastic model

In this subsection I discuss briefly a model with internal (multiplicative) Gaussian noise. The motivation is two-fold: First, the time series generated by the stochastic model (using parameters determined via the deterministic model in one of the cases discussed in the preceding subsections) furnish surrogate data to be analyzed using the same (deterministic) procedure applied to the observational data, as a means of estimating the uncertainties in the fit parameters *β*_2_, *β*_3_, *t*_1_ and *t*_2_. Second, the results of stochastic simulations allow one to judge the extent to which irregularities in the observational time series can in fact be attributed to internal fluctuations.

The equations governing the stochastic model are:

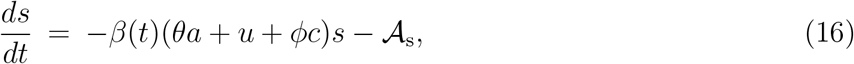

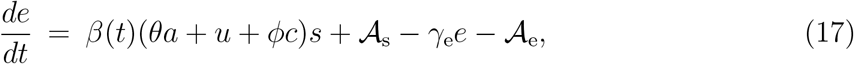

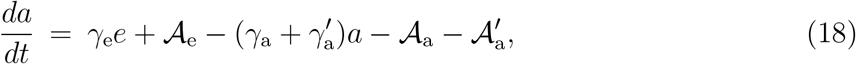

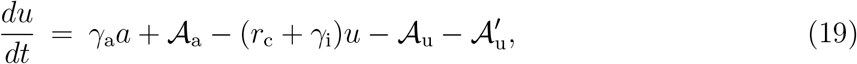

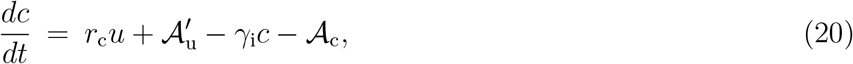

The cumulative case number follows,

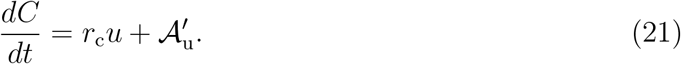

The noise term 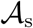 is defined so,

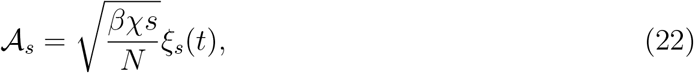

where *χ* = *θa* + *u* + *ϕc* and *ξ*_s_(*t*) is zero-mean, unit-intensity, Gaussian white noise, i.e., (*ξ*_s_(*t*)*ξ*_s_(*t*′)) = *δ(t − t*′). The other noise terms are defined analogously; the noise terms are mutually independent.

The set of stochastic differential equations are integrated numerically using an Euler scheme with timestep *h* = 0.01. One step in the evolution of *s* is given by,

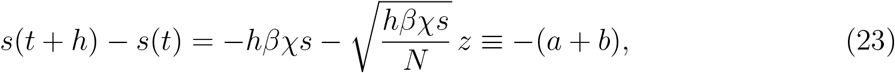

where *z* is a Gaussian random number with zero mean and unit variance, and *a* and *b* denote the deterministic and stochastic contributions, respectively. Analogous expressions hold for the other equations. It is essential that at each step, the population fraction *a* + *b* transferred from one state to another be nonnegative. This condition is enforced by “tempering” the noise [14, 15] such that *−a ≤ b ≤ α*. In the rare cases in which *a* + *b* < 0, it is reset to zero, and similarly, if *a* + *b* > 2a it is reset to 2*a*. (Note that tempering only on the negative side would induce a bias toward positive noise values.)

I compare observational data for the Brazilian state of Sergipe, the deterministic model fit, and twenty independent realizations of the stochastic model (using the fit parameters obtained from the deterministic model) in Fig. 10. The stochastic model yields a set of smooth curves which follow the deterministic evolution quite closely. Since the initial condition for the stochastic model is the same used in the deterministic analysis, the dispersion about the deterministic model is initially small, although it grows to encompass the fluctuations in the observational data in the latter part of the evolution. It is nevertheless clear that the stochastic model does not reproduce the fluctuations in the observational data, which is characterized by a number of small jumps and slowly varying oscillations (with a coherence time of roughly ten days) about the model fit. In other words, the observational fluctuations are not explained completely by internal fluctautions included in the stochastic model. Sources of additional fluctuations may include (1) independent outbreaks in localized regions beginning on different days; (2) changes in reporting policy or testing capacity; (3) variations in intervention policies between locales and over time.

**FIG. 10:**
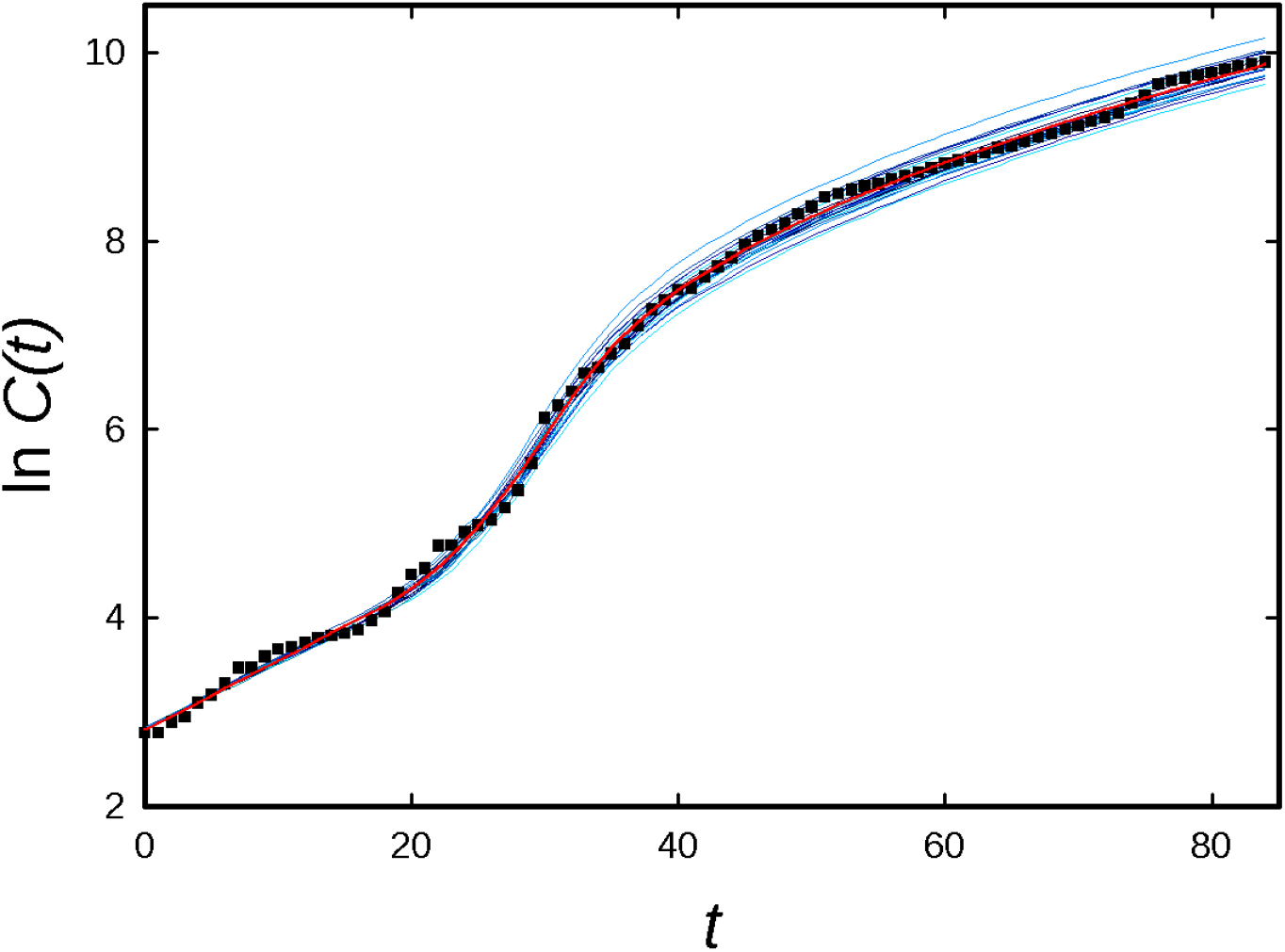
Observational data for the Brazilian state of Sergipe (points), model fit (thick red curve) and twenty realizations of the stochastic SEAUCR equations (thin blue curves) using the fit parameters obtained in the model fit.

Applying the same analysis used to fit the observational data to the stochastic time series, one finds minimal variations in parameters *β*_1_ and *β*_3_, and uncertainties (given by the standard deviation of the mean) of 0.05 in *β*_1_, and of 0.7 and 0.3 days, respectively, in *t*_1_ and *t*_2_. Analysis of stochastic time series for the case of Fairfield County, Connecticut yields similar conclusions regarding fluctuations and similar values for the uncertainties in the fit parameters (in this case the uncertainty in *β*_2_ is 0.03). The stochastic simulations yield the following estimates for the relative uncertainty in the final number of cumulative cases: 2.6% (Sergipe, 85th day of fit) and 1.0% (Fairfield, 89th day of fit).

### E. **Uncertainty estimates of parameters in** *β*(*t*)

The smoothness of the time-series generated using the stochastic model suggests that analyses of these series may underestimate the uncertainties in the parameters *β*_2_, *β*_3_, *t*_1_ and *t*_2_. (Additionally, analyzing sets of stochastic time series for each region treated in this study would be very time consuming.) As an alternative, I examine how the quality of fit, as reflected in the mean-square error (m.s.e.) defined in Eq. (15), changes as these parameters are varied. Denoting the best-fit parameter values by 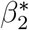, 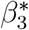, 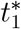 and 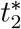, I determine the smallest change Δ*β*_2_ such that using 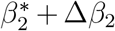 (with other parameters fixed at their best-fit values) causes the m.s.e. to double. Δ*β*_2_ is then taken as an uncertainty estimate for *β*_2_, and similarly for the other fit parameters. (While this procedure ignores possible compensatory effects when two or more parameters are varied simultaneously, it should yield preliminary uncertainty estimates.) I use the same resolutions in Δ*β_i_* (0.01), and in Δ*t_i_* (one day) as employed in the fitting procedure. The resulting estimates are given in Table I in the form of means and standard deviations for the three data sets analyzed.

The average estimated uncertainties are approximately one to two times the parameter resolutions. The relatively large standard deviation in Δ*β*_2_ for the set of U.S. counties and cities is due principally to two cases in which Δ*β*_2_ = 0.20. Both are associated with a single-day interval (i.e., *t*_2_ = *t*_1_ + 1) such that varying *β*_2_ has minimal impact on the quality of fit. In all three data sets, there are many instances in which changing *β*_3_ by 0.01 results in a sizeable (five- or ten-fold) increase in the mean-square error, suggesting that a somewhat higher resolution could be used for this parameter.

**TABLE I:** Fit-parameter uncertainty estimates as described in text for three data sets. Each entry reports the average uncertainy over the set, followed in parentheses by the standard deviation.

| Set | $\Delta\beta_2$ | $\Delta\beta_3$ | $\Delta t_1$ | $\Delta t_2$ |
| --- | --- | --- | --- | --- |
| U.S. | 0.024 (0.044) | 0.012 (0.005) | 1.1 (0.5) | 2.1 (1.7) |
| Brazil | 0.016 (0.014) | 0.014 (0.006) | 1.1 (0.3) | 2.1 (1.3) |
| World | 0.018 (0.024) | 0.014 (0.010) | 1.2 (0.6) | 1.8 (1.6) |

## IV. DISCUSSION

In this section I examine some tendencies evident in the results.

### A. Evolution of contagion rates

As is evident in Fig. 1, there is a general (though not universal) tendency toward reduced infection rates. This is reflected in the progression of sample-average *β* values shown in Fig. 11. In all three samples, 〈*β*_3_〉 is the smallest; for the U.S. and World samples, the mean of *β*_2_ is substantially smaller than that of *β*_1_, while for Brazil, 〈*β*_2_〉 ≃ 〈*β*_1_〉, which is substantially smaller than 〈*β*_1_〉 for the other two sets. The inset of Fig. 1 shows a similar analysis pooling all three sets, grouped into 55 regions with population *N <* 5 × 10^6^, 48 with 5 × 10^6^ < *N* < *β* × 10^7^, and 35 with *N* > *β* × 10^7^; the tendency of the contagion parameter to decrease is if anything more evident in this case. In Fig. 12 the mean value of *β*(*t*) is plotted over time for the three data sets. For each region, time zero corresponds to the first day of the time series included in the fit. The overall reduction in the contagion parameter is evident, as well as the tendency to a smaller dispersion of values.

**FIG. 11:**
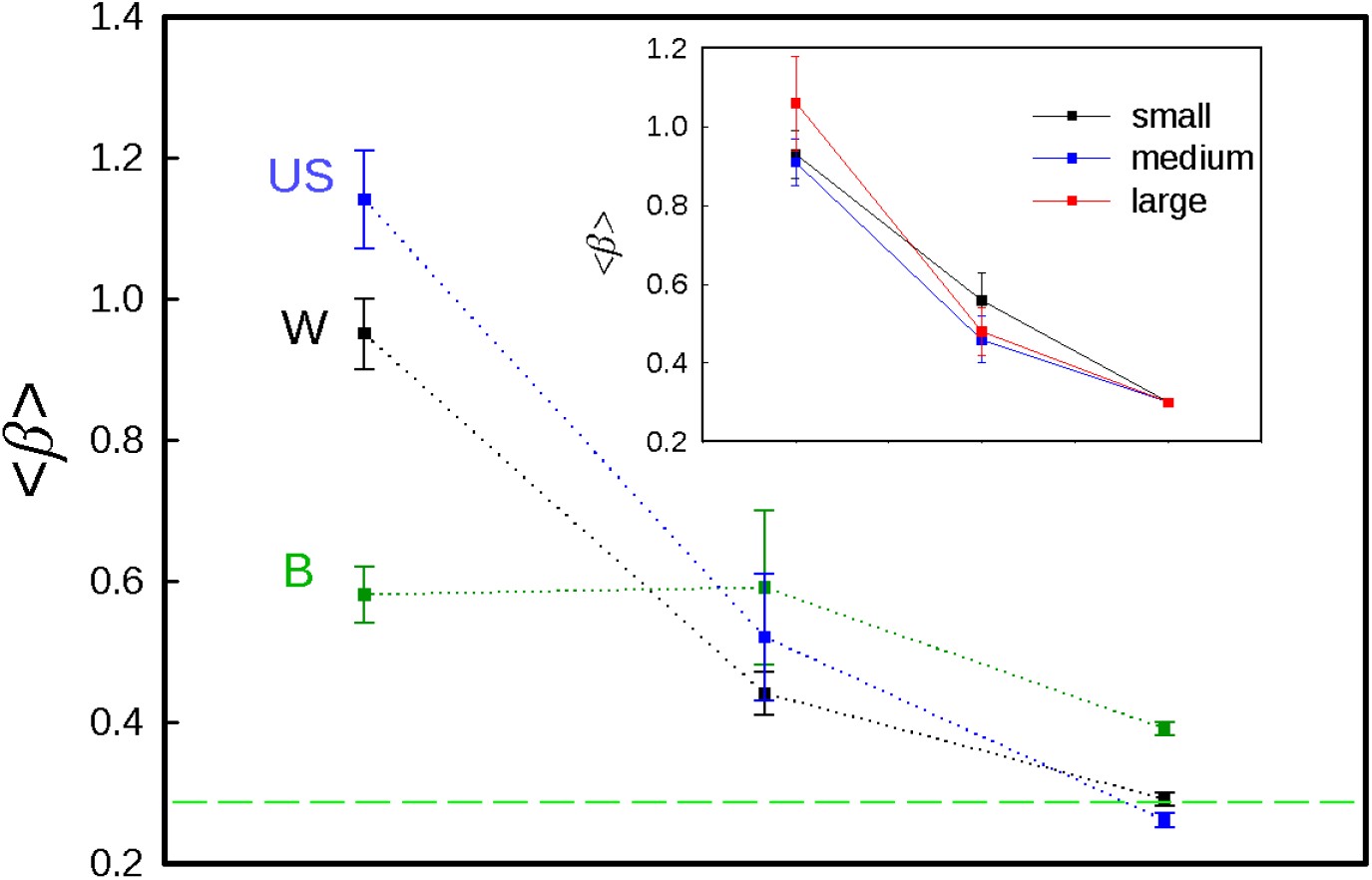
Mean values of *β*_1_*, β*_2_ and *β*_3_ for U.S. counties and cities, Brazilian states (B), and countries (W). The dashed horizontal line denotes the critical value, *β_c_*. Inset: averages for regions with small, medium and large populations as defined in text. Error bars denote standard deviations of the mean.

**FIG. 12:**
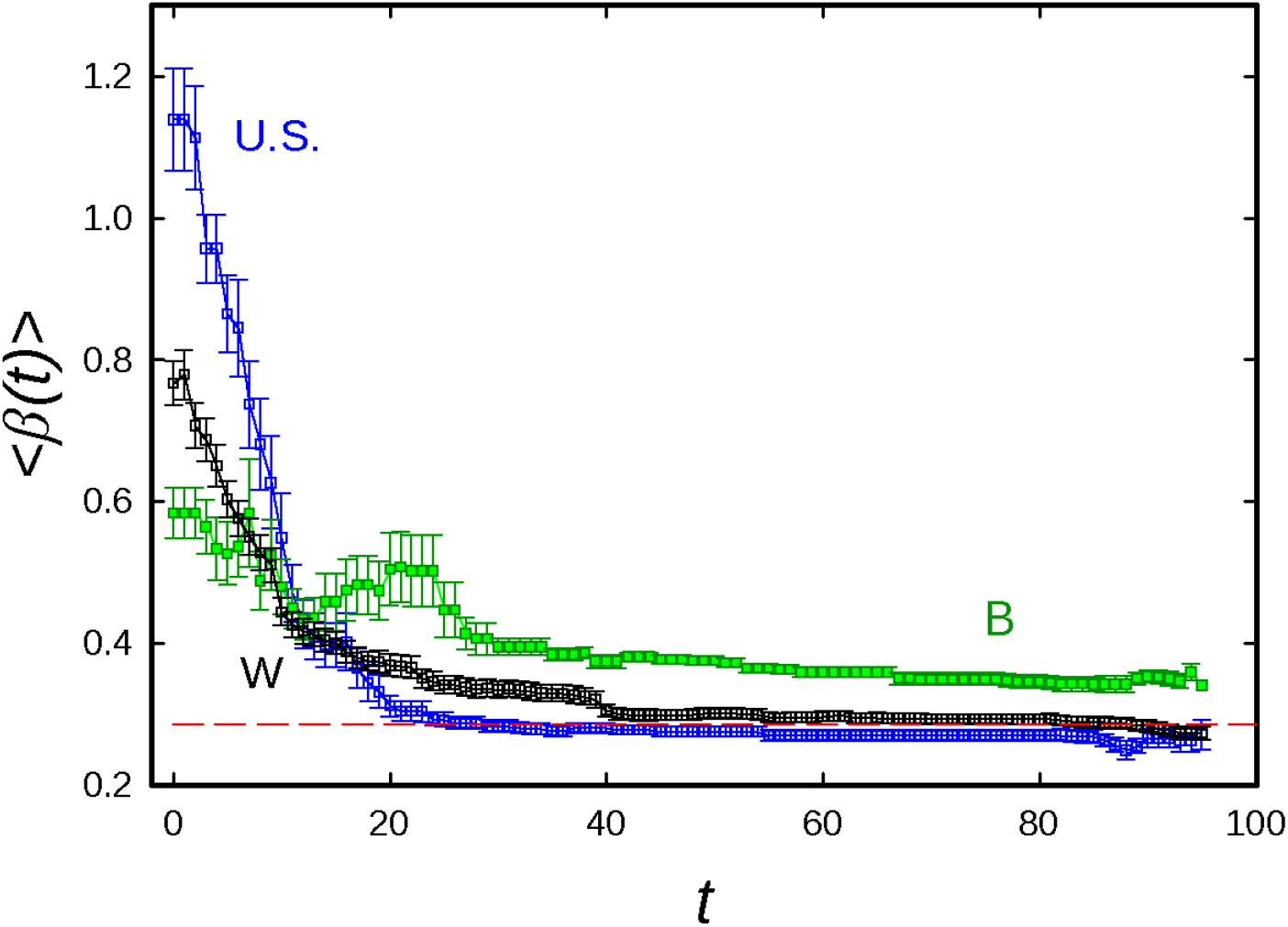
Mean value of *β* versus time for U.S. counties and cities, Brazilian states (B), and countries (W). Here time zero corresponds to the first day fit for each region. Error bars denote standard deviations of the mean. The dashed horizontal line denotes the critical value, *β_c_*.

### B. Sensitivity to changes in epidemiological parameters

In the studies reported above, the epidemiological parameters γ_e_, γ_a_, 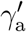, γ_i_, *r_c_* and *ϕ* are kept fixed at the values specified in Sec. II. Here I examine how some of the results, and the quality of fit, change under modest rescalings (reduction or increase by 20%) of each such parameter, maintaing the others fixed. Intuitively, reductions in γ_a_, 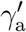, and γ_i_, prolong the mean lifetime of contagious states *a, u* and *c*, so that the observational data can be fit with smaller values of *β*; conversely, increases in these rates should be associated with increased *β*. Reducing *r_c_* can result in conflicting effects: First, slowing the transition from U to C (which is less contagious) should reduce *β*.; on the other hand, the reduction in confirmed cases (for the same number of infections) might require a larger a larger *β* to reproduce the observation case numbers. The effect of changes in *γ*_e_ on *β* is less obvious. By contrast, since the relative growth rate, 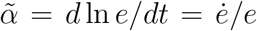, is strongly linked to the observational case numbers, one would expect an “ideal” model to yield predictions for 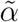 that are independent of parameter values. (To be more precise, the prediction for 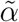 under *steady growth* should be insensitive to parameter variations; such variations do affect the response times, so that transients would in principle be affected.)

In Table II I report the changes in *β*_3_ and in 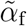 (the value on the final day of the series) provoked by 20% increases and reductions in the various parameters, as well as in the m.s.e. of the best fit, in the form of averages over the set of 103 countries analyzed above. (Using the original parameter set, the average m.s.e. is 2.8(3) × 10^−3^.) The changes induced in *β*_3_ follow the anticipated trends for γ_a_, 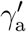, and γ_i_. For reasons that are unclear, the response to a change in *γ*_e_ is similar. Changes in 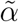 are generally less significant, relative to uncertainty, and do not follow a consistent pattern. The alterations induced by changing *r_c_* and *ϕ* are virtually insignificant. Finally, only changes in *γ*_e_ and *γ*_a_ lead to significant alterations in the quality of fit. The modest increases in m.s.e., both for increases and reductions, suggest that these parameters are well chosen. The absence of significant changes in m.s.e. in response to variations in the other parameters suggests that the values used here are not strongly in error.

**TABLE II:** Changes in *β*_3_, 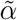, and the mean squared error (mse) induced by changes in a given parameter. In each entry, the first number corresponds to a 20% reduction in the parameter, the second to a 20% increase. Values are means over the set of 103 countries analyzed above; numbers in parentheses are uncertainties (standard deviation of the mean).

| Par. | $\langle \Delta \beta_3 \rangle$ | | $\langle \Delta \tilde{\alpha} \rangle (\times 10^{-3})$ | | $\langle \Delta(\text{m.s.e.}) \rangle (\times 10^{-4})$ | |
| --- | --- | --- | --- | --- | --- | --- |
| $\gamma_e$ | -0.023(3) | 0.030(3) | 5(2) | 3(2) | 3(2) | 6(3) |
| $\gamma_a$ | -0.008(3) | 0.013(4) | -3(4) | -1(5) | 4(2) | 4(2) |
| $\gamma'_a$ | -0.017(2) | 0.026(3) | 2(2) | 4(2) | 2(2) | 2(3) |
| $\gamma_i$ | -0.022(3) | 0.026(3) | 4(3) | 1(2) | 3(2) | 3(2) |
| $r_c$ | -0.002(2) | 0.006(2) | 1(2) | 2(2) | 2(2) | -1(1) |
| $\phi$ | -0.005(2) | -0.001(2) | 1.5(1.2) | 1.6(1.5) | 1(1) | 1(1) |

### C. Fraction of exposed individuals

A critical issue governing the evolution of the pandemic is the fraction of the population that has been exposed to the virus, since, in the absence of an effective vaccine or strict mitigation policies, only broad exposure and attendant immunity can halt the spread of infection. While the global (cumulative) number reported cases represents a small fraction of the world population, estimates of the prevalence of unreported infections (due to asymptomatic or weakly symptomatic cases, as well as to limitations in testing) suggest total case numbers larger than reported values by a factor of five, ten or even more. Subject to this uncertainty, the model employed here furnishes an estimate of the exposed fraction.

For the parameters employed in the present analysis, about four fifths of cases go unreported. The cumulative fraction of exposed individuals on thefinal day of the time series, *e*_tot_ = 1 − *x_f_*, ranges from < 0.1% to about 25%, with a strong tendency toward small fractions in highly populated regions. In particular, all regions with *e*_tot_ ≥ 0.1 have populations smaller than 2 × 10^7^; only six regions have *e*_tot_ > 0.2, and these have populations smaller than *β* × 10^6^. The general trend is illustrated in the scatter plot of Fig. 13, which shows that larger values of *e*_tot_ become increasingly rare as population size N increases. A population-weighted mean over the 103 countries studied yields a mean value of 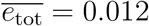. Although the restriction to regions with 2000 or more cases means that many regions are excluded, it is worth noting that the 103 included regions correspond to about 70% of the world population.

**FIG. 13:**
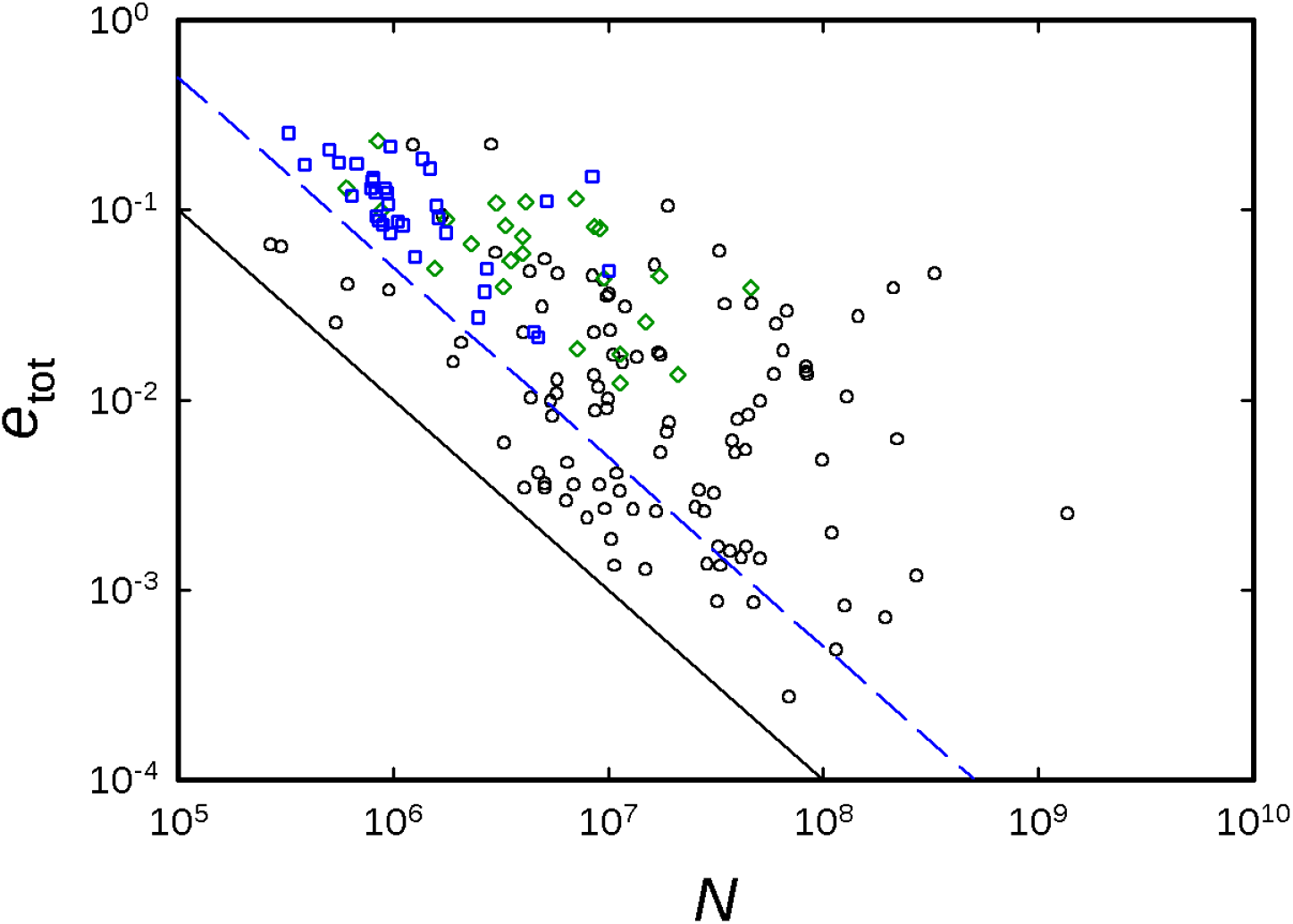
Cumulative exposed fraction *e*_tot_ versus population size for 103 countries (circles), 34 U.S. counties and cities (squares), and 24 Brazilian states (diamonds). The diagonal lines show the lower limits on *e*_tot_ imposed by the criteria of 2000 or more reported cases (countries and Brazilian states) and 10 000 or more cases (U.S.), recalling that the model assumes that actual case numbers are about five times greater than the reported values.

Two regions with rather high values of e_tot_ (as of 12 June 2020) are New York City and Suffolk County, Massachusetts, for which the model prediction is 15%. These tally reasonably well with antibody testing results of about 20% exposed in New York City (19 April 2020) [8] and 10% in Boston (15 May 2020) [16].

## V. CONCLUSIONS

I have shown how a simple SEIR-like model with a time-dependent infection rate, *β*(*t*), is capable of fitting many time series of confirmed Covid-19 cases. This supports the assertion that, at the present stage of the pandemic, reductions in the exponential growth rate are principally due to interventions (social isolation, lockdowns, mask-wearing) and not to depletion of the population of susceptibles. Even under the most “optimistic” scenarios, it is difficulat to imagine that the global fraction of the exposed population is more than a few percent (although it may be an order of magnitude greater in specific limited regions). The observational data and the model-based analysis both point to a strong suppression of contagion in many, but not all cases. Since the model lacks spatial structure, one would expect it to be most appropriate for relatively small regions such as cities, states or provinces, or smaller countries. A collection of such models could then be used to represent regions connected by movement of individuals [9, 10]. It is nevertheless interesting that the simple model captures the evolution in larger regions rather well.

Simulations of a stochastic version of the model permit one to estimate the contribution to uncertainties in *β*(*t*) due to intrinsic fluctuations, and show that in general, the latter do not explain the apparent noisyness of the data. A preliminary analysis suggests that *β* values are typically uncertain by a few percent, and switching times by a day or two. Changes in the quality of fit under varions of the epidemiological parameters (excluding *β*(*t*)) are quite modest, suggesting that the values adopted here are plausible, and sufficient for modelling on this level of detail.

Given the overall success in fitting observational data using a simple (piecewise constant) representation of *β*(*t*), the model proposed here seems a good candidate for continued studies. In analysing longer time series, additional time intervals and associated *β* values will be required, but this appears to be a straightforward extension of the current approach. Another short-term goal is to analyze mortality time series in parallel with case numbers, which should provide a test of model consistency as well as estimates of mortality rates. A more ambitious (and uncertain) goal is that of predicting the consequence of changes in intervention policies, a task that requires analysis of how such interventions have affected contagion up to now.

## Data Availability

All data are available as URLs.

https://raw.githubusercontent.com/CSSEGISandData/COVID-19/master/csse_covid_19_data/csse_covid_19_time_series/time_series_covid19_confirmed_global.csv

https://github.com/wcota/covid19br/blob/master/cases-brazil-states.csv

## Acknowledgments

I am grateful to Silvio Ferreira, Dean Karlen, Ricardo Takahashi, Gustavo Guerrero, Luiz Duczmal, Denise Bugarelli Duczmal, Armando Neves, Henrique Paiva, Ole Peters, Hans Herrmann and Constantino Tsallis for helpful discussions. I thank Leonardo Ferreira Calazans and Kevin Liu Rodrigues for valuable assistance in data gathering and visualization.

